# Evaluation of Measles Surveillance System amidst Covid – 19 pandemic in Asutifi North District, Ahafo Region, Ghana

**DOI:** 10.1101/2021.03.10.21253259

**Authors:** Sekyere Stephen Owusu, Laar Salam Dam-Park

## Abstract

**Background:** Measles is a disease of public health importance earmarked for elimination by all WHO Regions. Globally, more than 140 000 people died from Measles in 2018 affecting mostly children under 5 years, despite the availability of safe and effective vaccine.

**Methods:** A descriptive cross-sectional survey was conducted. Disease surveillance focal persons were interviewed using semi-structured questionnaire on the system operations and use of Measles case definitions. Measles case-based investigation forms from 2015 – 2020 were reviewed for its timeliness and data quality. CDC updated guidelines for surveillance system evaluation was used to assess its usefulness and attributes. Data was analyzed for frequencies and proportions and results presented in tables and graphs.

**Results:** Measles surveillance system was timely as 100% (69/69) of the suspected cases were reported on time. Also, the level of representativeness was good as all the 14 health facilities in the District were participating in the Measles Surveillance system. Majority 73.1 (44/60) of the case-based investigation forms filled were incomplete with some columns wrongly filled.

**Conclusion:** Despite the outbreak of Covid – 19 with most districts battling with how to contain the virus, measles surveillance system was still meeting its objectives of early detection and prompt reporting but with poor data quality.

## INTRODUCTION

Measles is an acute viral infectious disease and an important cause of childhood morbidity and mortality worldwide [1]. Measles is transmitted mainly through direct contact (coughing and sneezing) and also by small-particle aerosols in an airspace where an infected person has coughed or sneezed in the previous few hours[2].Typically, with prodromal symptoms including cough, coryza, and conjunctivitis[3]. There is no specific antiviral treatment. In addition, Measles infections cause long-lasting memory B and T cell impairment, predisposing people susceptible to opportunistic infections for years[4].Exposed people who are not immune have up to a 90% chance of contracting the disease, and each person with Measles may go on to infect 9 to 18 others in a susceptible population[5]. Laboratory confirmation of Measles virus infection can be based on a positive serological test for Measles-specific immunoglobulin M antibody[6]. Measles can be occurred in all age groups. However, children younger than 5 years of age and adults older than 20 years of age are more likely to suffer from Measles complications[7]. Promoting vitamin A supplementation in children with Measles contributes to the control of blindness in children[8]

More than 140 000 people died from Measles in 2018 mostly children under the age of 5 years[9]. Despite the existence of a safe and effective vaccine, Measles remains a major cause of morbidity and mortality globally, especially in young children [10]. Overall, 176,785 confirmed Measles cases were reported in African Region(AFR) through case-based surveillance during 2013–2016 [11]

The 47 Member States of the AFR of the World Health Organization established a goal in 2011 to achieve Measles elimination by 2020 using the following strategies: attaining high routine immunization coverage; conducting Measles supplemental immunization activities (SIAs); conducting case based surveillance with laboratory confirmation of suspected cases and improving management of Measles cases[12]

In Ghana, about 124 Measles cases were recorded in the year 2014 but as at 2018, there was a significant decrease to about 34 Measles cases. Measles incidence and mortality rates have significantly decreased since vaccine introduction[13]. Despite substantial residual susceptibility among young adults, more in some locals than others, sustained routine childhood immunization likely would eliminate Measles eventually [14]. Measles remains a major public health problem in many developing countries in which vaccination coverage is poor[15].

Notwithstanding, since the first case of a novel coronavirus (COVID-19) infection was detected in Wuhan, China on 31^st^ December, 2019, a series of confirmed cases of the disease has been recorded all over the world. The level of global spread and severity of the disease lead the World Health Organization (WHO) to characterize the disease as a pandemic on 11^th^ March, 2020. This has halted a lot of activities with most organizations shifting their attention to the preventive measures of containing the outbreak and preventing further spread. As at the time of data collection, Asutifi North district has recorded 129 confirmed positive cases of Covid – 19.

We evaluated the Measles Surveillance System in Asutifi North district to assess its attributes, usefulness and objectives in the mist of Covid – 19 pandemic.

## METHODS

### Study design

We conducted a descriptive cross-sectional study in Asutifi North District. We used Centers for Disease Control and Prevention (CDC) updated guidelines for evaluating public health surveillance systems 2001.

### Study area

We conducted the study in Asutifi North District of Ahafo Region, Ghana. The study area has a projected population of 67206. The District is divided into four Sub-Districts. In relation to health care delivery, the District has 9 government health facilities with one private hospital, one private maternity home and three private clinics. Seven out of 14 health facilities in the District has laboratory capacity to collect appropriate Measles samples when a case is suspected in the facility.

### Study population

Participants included Surveillance Officer/ focal persons, Laboratory Technicians and Community Based Surveillance Volunteers involved in Measles surveillance.

### Data Collection

Convenient sampling method was used to sample respondents from each health facility in the district. We interviewed participants to determine their knowledge on the operations of the surveillance system and assess their attributes. Completed Measles case based investigation forms from January 2015 up to December 2020 were reviewed to check for systems attributes like data quality, simplicity, completeness, sensitivity, predictive value positive and timeliness of the system.

### Measles case definition

#### Suspected Measles

Any person with fever and maculopapular (non-vssicular) generalized rash and cough, coryza or conjunctivitis (red eyes) or any person whom a clinician suspects Measles.

#### Confirmed Measles

Any suspected case with laboratory confirmation (positive IgM antibody) or epidemiological link to confirmed cases in an outbreak.

### Data Analysis

Measles data was captured using Epi Info Version 7 and analyzed with Microsoft Excel to generate frequencies, tables and graphs.

### Ethical considerations

Permission to carry out the study was sought from the Ahafo Regional Health Directorate and Asutifi North District Health Directorate. Confidentiality was assured and maintained throughout the study period.

### Assessment of Attributes and Usefulness of Measles Surveillance System

#### System usefulness

We assessed it based on its objective of detecting Measles cases and its contribution towards disease prevention.

### Qualitative attributes

#### Simplicity

It was assessed on time spent on collecting data

#### Flexibility

It was assessed retrospectively by observing how the system has responded to a new demand.

#### Acceptability

We assessed acceptability by interviewing participants on their willingness to participant in the surveillance system.

#### Representativeness

We assessed it on system ability to accurately describes the occurrence of Measles cases over time and its distribution in the population by person and place

### Quantitative attributes

#### Sensitivity

We assessed it on the number of Measles cases detected by the surveillance system

#### Data quality

It was assessed by reviewing the Measles reports to examine wrong entries and incompleteness of data.

#### Timeliness

We assessed the speed between steps in the Measles surveillance system usually from the onset of the health event and it reporting to the next level.

#### Stability

We assessed on the system’s ability to collect, manage and provide data without system failure.

## RESULTS

### Data Reporting

Measles cases when suspected at the community level by Community Health Officer or Health Worker or a CBSVs, the case is referred to the nearest health facility within the Sub-district for sample collection. Also, when a case is been suspected at the facility level, appropriate sample is taken and sent to District Health Directorate for onward submission to Region Health Directorate Public Health Unit.

At the regional level, the sample is received and sent to National Public Health Reference laboratory for confirmation. Laboratory results at the Public Health Reference laboratory are sent to the National Public Health Unit. From the National Public Health Unit, data on laboratory confirmation are sent to WHO and other partners. National Public Health Unit upon receiving the laboratory results, will timely send feedback to Regional Public Health Unit for submission to Districts, facilities and patients.

This study found an increasing trend of Suspected Measles cases in Asutifi North from 2015 to 2019 and a sharp decline in the year 2020 (Figure 3).

**Figure 1:**
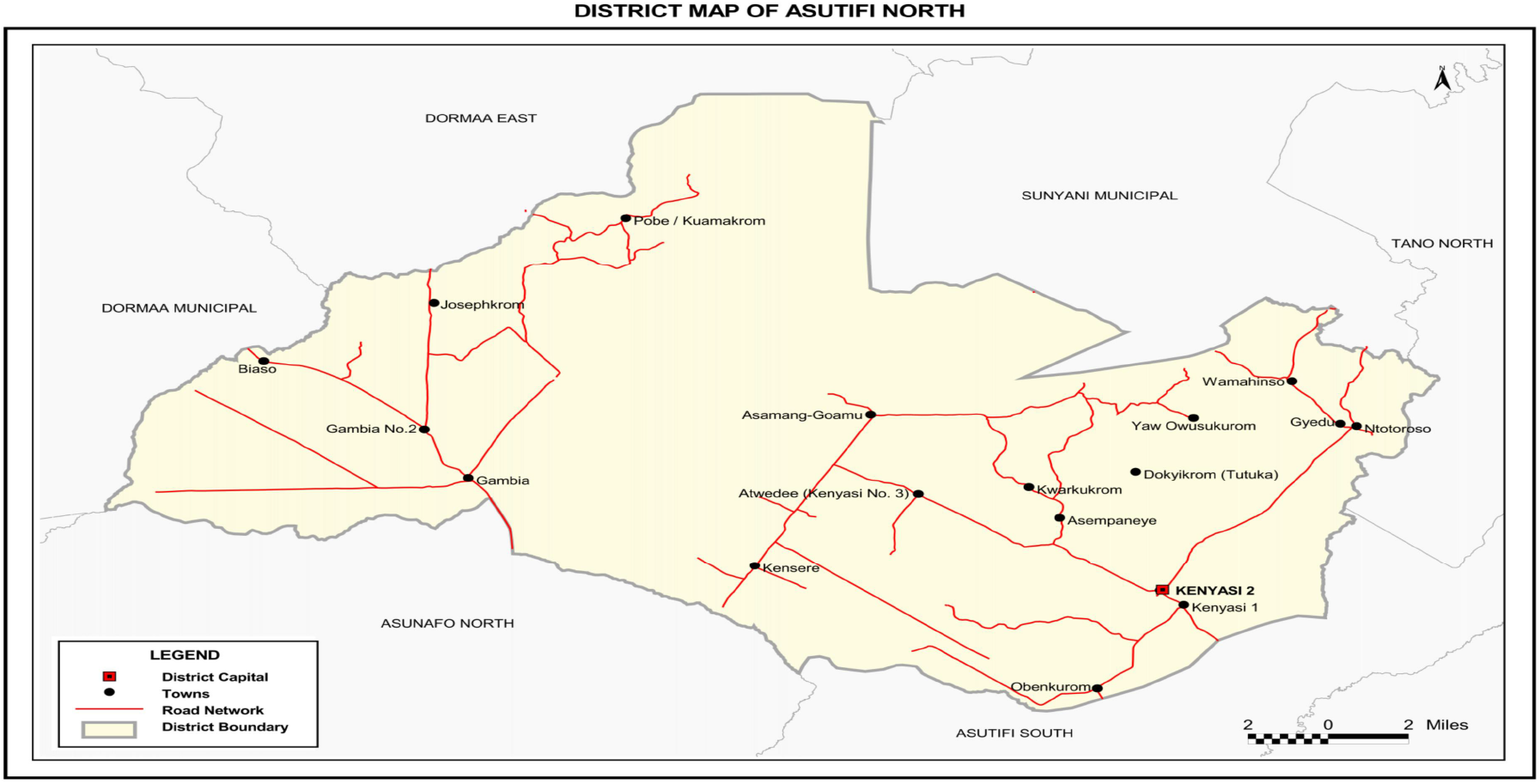
Asutifi North District map (Source District Health Directorate)

**Figure 2:**
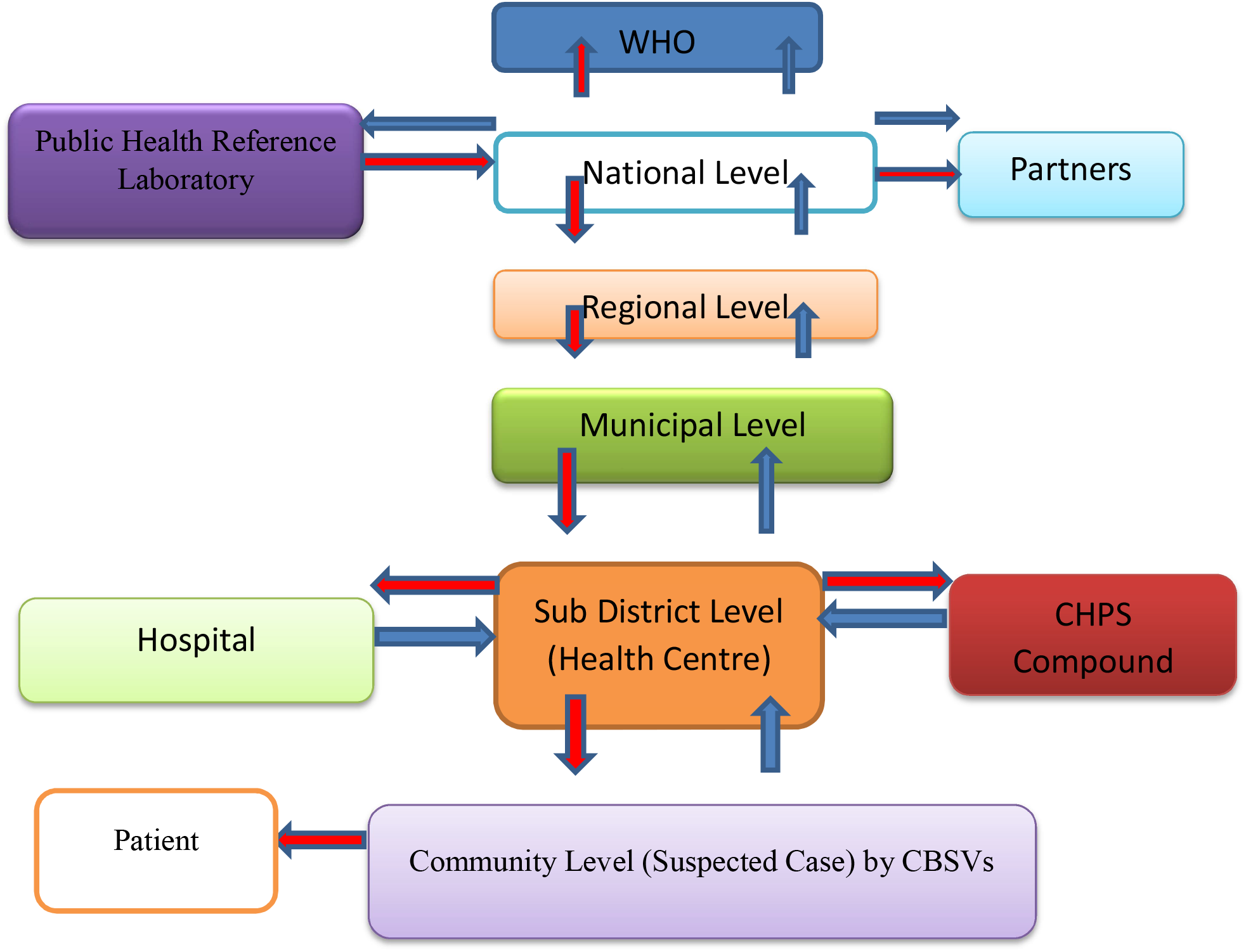
Measles Surveillance System Flow Chart.

**Figure 3:**
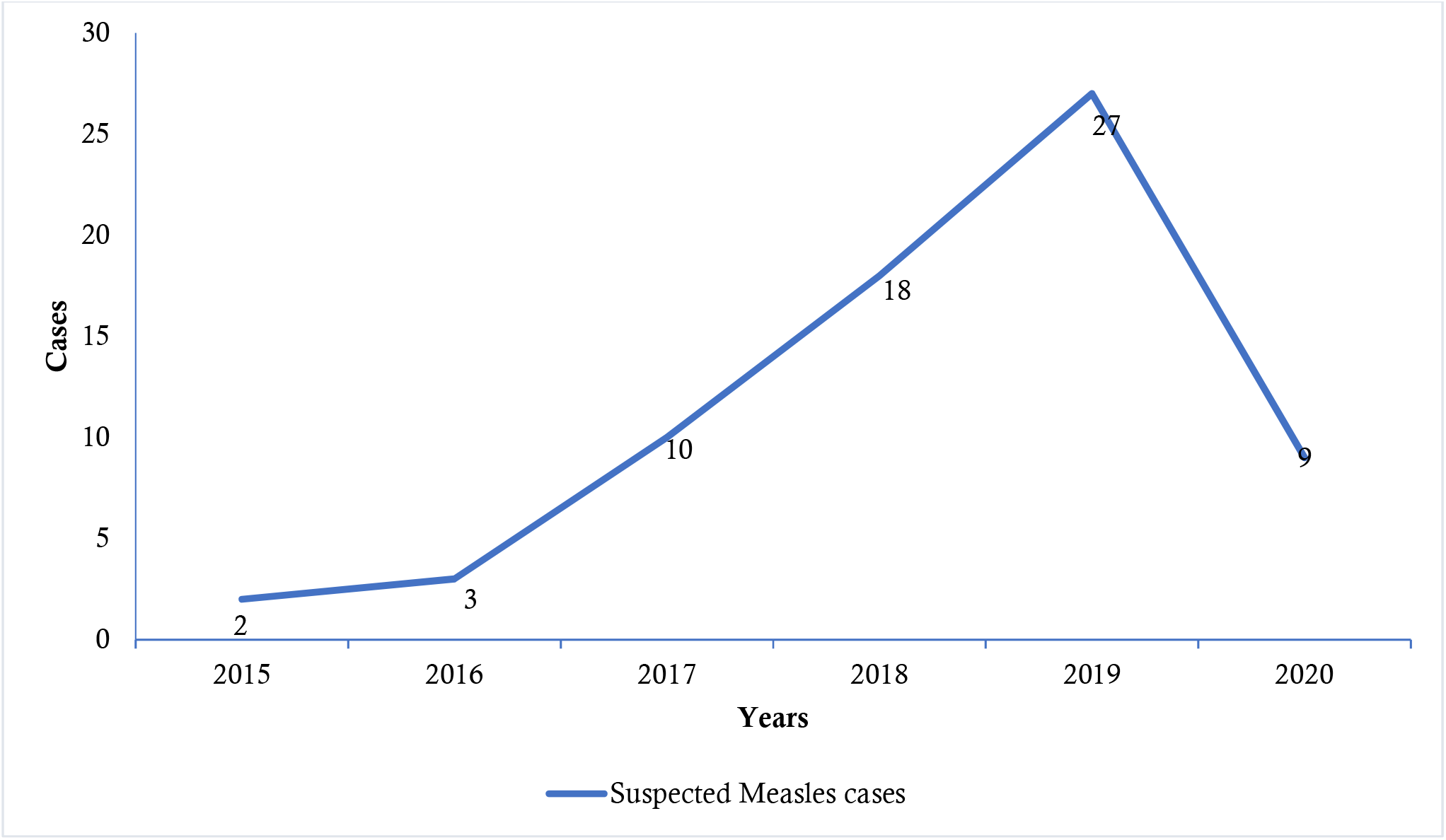
Trend of suspected Measles cases in Asutifi North 2015-2020.

### Objectives and usefulness of the Measles Surveillance System

The Measles surveillance system was useful. Data generated provided basis for some public health actions such as 2018 Measles and Rubella vaccination campaign in the District. The district also received two motor bikes to support surveillance activities in 2019. The surveillance system has achieved it objectives by detecting and responding promptly to cases of Measles. Total of 69 suspected Measles cases were detected from 2015 to 2020.

### Measles Surveillance System Attributes

#### Simplicity

Majority, 86.7% (26/30) of the respondents said the surveillance system is simple since they require less time to fill Measles case-based investigation form (Table 1)

**Table 1:**
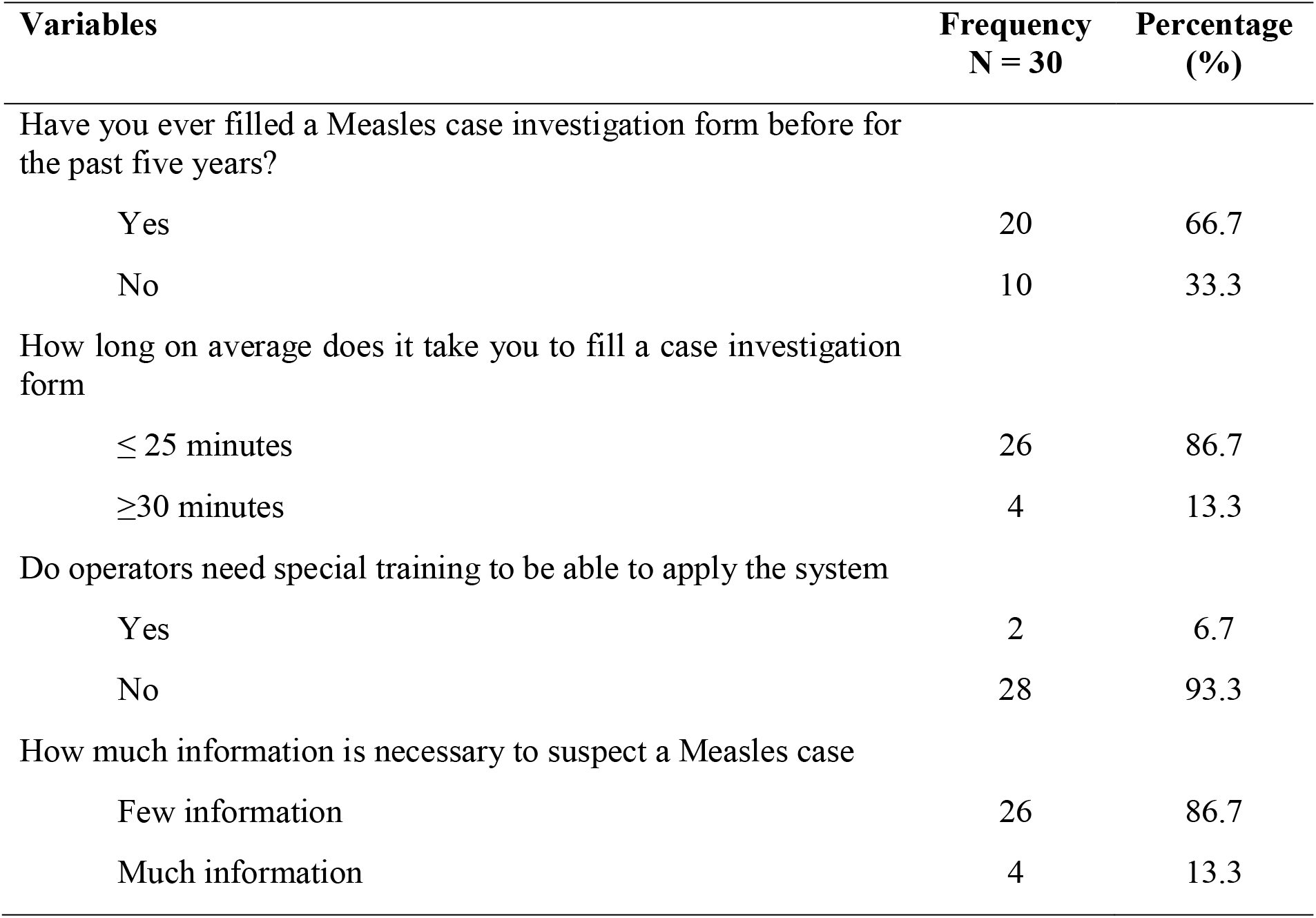
Simplicity of Measles Surveillance System in Asutifi North district, 2015-2020.

#### Flexibility

Majority 96.7% (29/30) of the respondents interviewed said that, Measles has been integrated into other surveillance systems. It was also observed that, there have been some changes in the system specifically on the case investigation form and the addition of oral swab as a sample for laboratory testing. The system was observed to have the ability to adapt to demanding changes.

#### Acceptability

Majority of the respondents 93.3% (28/30) felt the system was acceptable and said that it was their duty to fill Measles case-based investigation form. Also, all 30 respondents were willing to continue participating in the system.

#### Representativeness

The level of the representativeness was good as all the 14 health facilities including those from the private health facilities in the District were involved in the Measles surveillance system. Measles data captured information on socio-demographics, outcomes and geographical location from all reporting facilities.

#### Sensitivity

The system has been able to detect 69 suspected Measles cases from 2015 – 2020.The district has not recorded any Measles outbreak.

#### Data quality

Majority, 73.1% (44/60) of the forms reviewed over the 6year period were observed to be incomplete with some columns wrongly filled.

#### Timeliness

All (69/69) suspected cases of measles for the period under study (2015 −2020) were reported on time within 24hours of notification to the next level.

#### Stability

Staff and computers were available to operate Measles surveillance without any system failure. Cell phones are being used by the various facilities for reporting. Motor bikes were available for sample transportation.

## DISCUSSION

Periodic evaluation of the Measles surveillance system is essential in assessing the surveillance system attributes, usefulness and whether it is achieving it objectives. The Measles surveillance system was useful since data generated provided basis for some key public health actions such as 2018 Measles Rubella vaccination campaign and two motor bikes received in 2019 to support this surveillance system in the District

This study found Measles surveillance system in the District being sensitive for detecting 69 suspected cases from 2015 to 2020. There was no outbreak during this period.

Measles surveillance was found to be simple. Majority of the stakeholders said less time was required to fill a Measles case-based investigation form. This is supported by a study in Ethiopia [1] which reported that amount of time spent in collecting Measles surveillance data was less than 20 minutes.

Measles surveillance system had poor data quality with consistent incomplete case-based investigation forms and some columns wrongly filled. Our finding is similar to what was reported in Nigeria [21] which found the surveillance having a progressive decline in completeness of forms. This same finding is consistent in most part of Africa especially where paper-based reporting is used [22, 23].

The study found the Measles surveillance system to be flexible and simple. This is consistent with a similar study in Ethiopia [1] which reported that the Measles surveillance system is simple and flexible.

### Conclusion

Even though, the surveillance system evaluated was not excellent, it was generally good as it was able to suspect 69 Measles cases from 2015 to 2020. However, major challenges with regard to this surveillance system were poor data quality and lack of laboratory feedback of results. The effect of Covid – 19 pandemic could however been a factor that influenced the sharp decline of the number of suspected measles cases from 27 to 9cases between 2019 and 2020.

### Limitation

The major limitation was interviewees recalling events over the past five years, which was likely to be a recall bias, however, this was minimized since key surveillance actors were interviewed.

### Recommendation

- The District Disease Surveillance should organize orientation workshop for both key actors and Staff in the Measles surveillance system in filling of Measles case investigation form.
- District Director of the Health Services should liaise with Regional Deputy Director of Public Health for timely and regular feedback of suspected Measles cases sent for laboratory confirmation.

### Public Health Actions

We organized Measles Surveillance system orientation training for the District Disease Surveillance Officer, Facility Surveillance focal persons and Clinicians at both public and private health facilities. The orientation meeting was to enhance Measles case detection and ensure data quality for action and decision making in the District. Findings from the study was disseminated during District Health Management Team (DHMT) mid-year performance meeting.

## Data Availability

All data referred to in this article is readily available

## Acknowledgement

We thank the Regional and District Director for Ahafo region and Asutifi North District respectively for their support and permission granted for this output.Many thanks to Disease surveillance focal persons for their support.

